# Integrating Physiotherapy into Primary Care Models: A Scoping Review Protocol

**DOI:** 10.1101/2024.07.18.24310629

**Authors:** Nathaniel Saddy, Aamir Aboosally, Jordan Aslanidis, Anthony Beilin, Jessica da Silva Schezar, Jordan Miller, Andrews K Tawiah

## Abstract

**Introduction:** Physiotherapists (PTs) working in primary care settings within an interprofessional team can lead to favourable health outcomes and decreased burden on the healthcare system. Although PT models of care are important to primary care delivery, there is a lack of knowledge and evidence on the characteristics of these models of care, the differences and similarities between the models, and the barriers and facilitators to implementing these models. This scoping review protocol aims to fill this knowledge gap by synthesizing the evidence and characteristics of models of care that integrate physiotherapists within primary care teams, mapping the similarities and differences, and describing barriers and facilitators to implementing models of care that integrate physiotherapists within primary care teams.

**Methods:** The scoping review is based on the Joanne Briggs Institute (JBI) framework. It is reported according to the Preferred Reporting Items for Systematic Reviews and Meta-Analyses extension for Scoping Reviews (PRIMSA-ScR). A comprehensive search strategy will be used to find relevant papers in six databases: OVID MEDLINE, PubMed, Embase, CINAHL, Web of Science, and Scopus. Grey literature will be searched through OpenGrey, Theses Global, ProQuest Dissertation, and Google Scholar. Quantitative and qualitative study designs will be included, with two reviewers independently selecting each article on Covidence. Data will be extracted using a pre-piloted data extraction sheet and synthesized narratively to identify themes and patterns.

**Discussion:** This scoping review will synthesize the evidence on models of care that integrate physiotherapists within primary care teams. It will provide evidence to inform the implementation of these models of care and identify research gaps that need to be addressed.

## Introduction

According to the World Health Organization, primary health care is a whole of society approach that aims to maximize the level and distribution of health and well-being [1]. Primary Health care comprises three components 1) primary care and essential public health functions as the core of integrated health services 2) multisectoral policy and action 3) empowered people and communities. Primary care models are usually described as the first-contact service for a patient, and the models are built on an interprofessional holistic approach aiming for patient-centred care [2]. In a seminal article by Barbara Starfield, primary care is described based on four attributes known as the “four C’s”: First contact, Continuity of care, Comprehensiveness, and Coordination of care [3, 4]. These attributes have been expanded upon through a multi-stakeholder consensus process to formulate seven shared principles of primary care: 1) Person centered 2) Continuous 3) Comprehensive and equitable 4) Team-based and collaborative 5) Coordinated and integrated 6) Accessible 7) High value care [5]. There is evidence that strong primary care, regardless of whether it is delivered by a primary care physician or a team of health professionals, can prevent illness, improve health outcomes, and promote a more equitable distribution of healthcare to the population compared to a specialty care model [6].

Physiotherapists (PTs) can prevent, assess, and treat injuries, pain, diseases or disorders and their impact on function, movement, and overall health [7]. PTs are competent to practice independently and as part of an interprofessional team from primary to tertiary care. These competencies allow PTs to effectively be part of a team-based approach, an ability that is essential in primary health care models focusing on patient-centred care. Evidence suggests that multiple PT interventions are beneficial for treating multi-system conditions often seen in primary care. PTs have in-depth knowledge of musculoskeletal conditions, chronic pain management, and cardiorespiratory and neurological conditions, and these conditions are the areas of practice most commonly reported by PTs working in primary care [8]. PT services have demonstrated effectiveness in supporting people with several chronic conditions, including providing self-management support, activity counselling, and exercise prescription [8]. Recent studies have been conducted in Canada and Sweden to investigate PT models in primary care in specific health conditions and settings, such as orthopedic and low-back pain [9–11]. These studies found that PT models in primary care provided equal or better outcomes than usual care in primary care settings. However, these studies focused only on musculoskeletal (MSK) disorders.

The call for PTs in primary care has gotten more robust due to the challenges with access to primary care in multiple countries. Contributors to the lack of primary care access include staff shortages, service inequality by geographic regions, inefficient models of care that require referrals for certain primary care professionals, preparedness of healthcare providers, practitioner workload, and financial constraints [12]. In Canada, over 6.5 million people do not have access to primary care providers; this is 14.4% of the Canadian population [13], which is expected to grow. This has led to calls for interprofessional team-based models of primary care with physiotherapists as core members of the team. However, there is a lack of synthesized evidence on the models of care that integrate physiotherapists within primary care teams.

This scoping review protocol aims to fill this evidence gap by examining the evidence on the primary care models that integrate physiotherapists that have been implemented worldwide. This scoping review will identify and describe the characteristics of the models of care, mapping out the similarities and differences between the models. Further, this scoping review will identify the types of evidence available on models of primary care that integrate physiotherapists, which will help identify where evidence exists to inform implementation and where important research gaps need to be addressed. Finally, it will synthesize evidence barriers and facilitators to the implementation of primary care models that involve the integration of physiotherapists within primary care models to provide essential evidence to support the development of implementation processes for team-based models of primary care that include physiotherapists.

### Review questions

The specific review questions for this scoping review are:

1. What are the types of evidence available and gaps in evidence available related to primary care models that integrate physiotherapists within primary care teams?
2. What are the characteristics of primary care models that integrate physiotherapists and what are the similarities and differences between these primary care models?
3. What are the barriers and facilitators to implementing primary care delivery models that integrate physiotherapists within primary care teams?

## Methods

This scoping review will follow the Joanna Briggs Institute (JBI) framework for scoping reviews [14], following updated methodological guidance for the conduct of scoping reviews, best practice guidance and reporting for the development of scoping review protocols and recommendations for the extraction, analysis of presentation of results in scoping reviews [15,16]. The protocol is reported in alignment with the Preferred Reporting Items for Systematic Reviews and Meta-Analyses extension for Scoping Reviews (PRISMA-ScR) [17]. The protocol is registered on Open Science Framework registries at https://osf.io/kh83r/.

### Inclusion Criteria

The inclusion and exclusion criteria were designed based on the participants, concept, and context (PCC) framework described by the JBI Scoping Methodology Group [16]. Given that the focus of this scoping review is on models of health service delivery, participants were determined to be not a relevant inclusion criterion. The inclusion and exclusion criteria for this scoping review are presented in Table 1. The inclusion and exclusion criteria were designed to identify research related to the *concept* of interest (models of primary care that integrate physiotherapists within primary care teams) and the *context* (primary care settings anywhere in the world).

**Table 1.** Inclusion and Exclusion Criteria.

| Inclusion Criteria | Exclusion Criteria | Rationale |
| --- | --- | --- |
| The included studies will provide evidence of a model of care. As defined by the World Health | Studies that do not provide evidence on a model of care would be excluded. For example, research carried out | These inclusion and exclusion criteria will ensure the included studies are focused on **models of care**. |
| Organization (WHO), a model of care is “a conceptualization and operationalization of how services are delivered, including the processes of care, organization of providers and management of services, supported by the identification of roles and responsibilities of different platforms and providers along the pathways of care [18].” | in a primary care setting but aiming to assess the outcomes of a specific intervention (e.g., a medication) rather than a model of care would be excluded. A second example is research carried out in primary care that focuses on testing the psychometric or diagnostic properties of an assessment tool for a specific condition, which would be excluded. |  |
| A model of **primary care** (public or private). Primary care will be defined using Epperly’s shared principles of primary care, with emphasis on the following principles [5]:<br>(1) continuous (or longitudinal) care | Studies of a model of care that does not meet the definition or shared principles of primary care identified. For example, an emergency department does not typically offer longitudinal or comprehensive care, although it often includes interprofessional teams and is | These inclusion and exclusion criteria will ensure the studies included are focused on models of **primary care**. |
| (2) comprehensive care (the place in the health system where the average person turns for the majority of their routine healthcare needs)<br>(3) Team-based and collaborative care (at least one other health profession involved in addition to the physiotherapist)<br>(4) Accessible care (a setting where healthcare can be accessed directly without referral) | accessible as a ‘first-contact’ setting in most health systems.<br>A private physiotherapy clinic may provide direct access and sometimes longitudinal care but does not often offer comprehensive health services that meet most of the average person’s health needs. |  |
| Included studies will provide evidence on a primary care model | Studies on primary care models that do not include a physiotherapist. | These inclusion and exclusion criteria will ensure the studies included involve the |
| involving a licensed or registered **physiotherapist**. |  | integration of a **physiotherapist** within a primary care team. |

### Information Sources

A literature search will be completed across six databases: OVID MEDLINE, PubMed, Embase, CINAHL, Web of Science, and Scopus. Grey literature will also be searched through Web of Science, OpenGrey, Theses Global, ProQuest Dissertation, Google Scholar, and central physiotherapy professional and organizational websites. Quantitative and qualitative study designs, primary research, and systematic reviews will all be included in the scoping review.

## Data Management

Articles identified by the search will be imported and stored in COVIDENCE. The program will automatically remove duplicates. Title/abstract and full-text screening will be done through COVIDENCE, and full texts will be stored within the program.

### Search Strategy

The search strategy aims to identify published and unpublished grey literature with findings that will help further our understanding of the evidence on primary care models of health service delivery that involve the integration of physiotherapists within primary care teams worldwide. One research librarian within the Health Sciences Faculty at Queens University was consulted to assist with the creation of this search strategy. A preliminary search was conducted on OVID MEDLINE using our *concept* and *context*: “Primary Care” and “Physiotherapist” as key terms, along with relevant synonyms. We conceptualize “primary care” as the setting and processes in the health system that supports first-contact, accessible, continued, comprehensive and coordinated patient-focused care [1,3,5]. For the purposes of our inclusion criteria, we differentiate this from primary health care, which is a whole-of-society approach to health that aims to maximize the level and distribution of health and well-being through three components: primary care, essential public health functions, policy and action, and empower people and communities [1]. However, we acknowledge that these terms are often used synonymously in the literature and, therefore, have included “primary health care” as a synonymous search term to ensure we don’t miss relevant articles. Additional terms such as “First Contact” and “Advanced Practice” were incorporated to capture different representations of the “Primary Care” concept. Additional keywords were derived from the abstracts and titles of relevant studies from this preliminary search. These keywords are listed below in Table 2. Using these keywords and medical subject headings, six databases will be searched: OVID MEDLINE, PubMed, Embase, CINAHL, Web of Science, and Scopus. The search strategy will be modified for each unique database.

**Table 2.** Keywords used in the search strategy.

| **“Physiotherapy” descriptors** | **“Primary Care” descriptors** |
| --- | --- |
| Physiotherap*<br>Physical Therap* | Primary care, Primary health care, Primary Medical Care, Primary Care Model, Patient Care Team, Family Health Team, community health center, patient medical home, patient’s medical home patient centered medical home, health home |

All relevant literature published between 2003-Current in English and French will be considered in our search. These criteria were chosen based on the rationale of including the two main languages in the research team’s residing country (Canada) and attempting to include a broad range of studies that accommodate the team’s resources. The date criteria were selected because primary care models have undergone significant reform within the last decade in Canada and worldwide [18]. Therefore, the literature before 2003 may be scarcer and less relevant than recent studies.

A sample search strategy is provided in Table 3 below. As per the JBI search strategy recommendations, the reference lists of all identified articles will be screened to find any additional studies related to the research question. Finally, a grey literature search will also be conducted to capture information about our research question that is not represented in existing publications. Databases such as Web of Science, OpenGrey, Theses Global, ProQuest Dissertation, and Google Scholar will all be searched using a combination of the mentioned keywords. Finally, the websites of major professional and national physiotherapy organizations will be explored for further documentation of PT models of care in primary health settings.

**Table 3.**
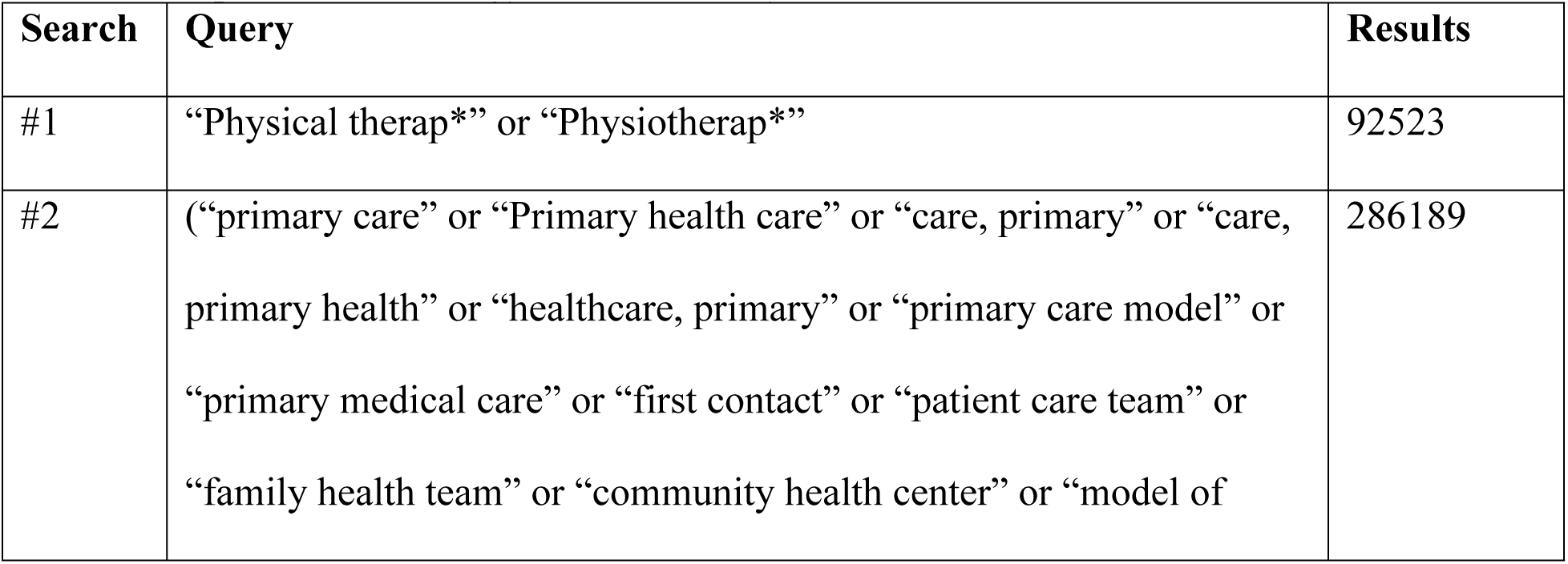
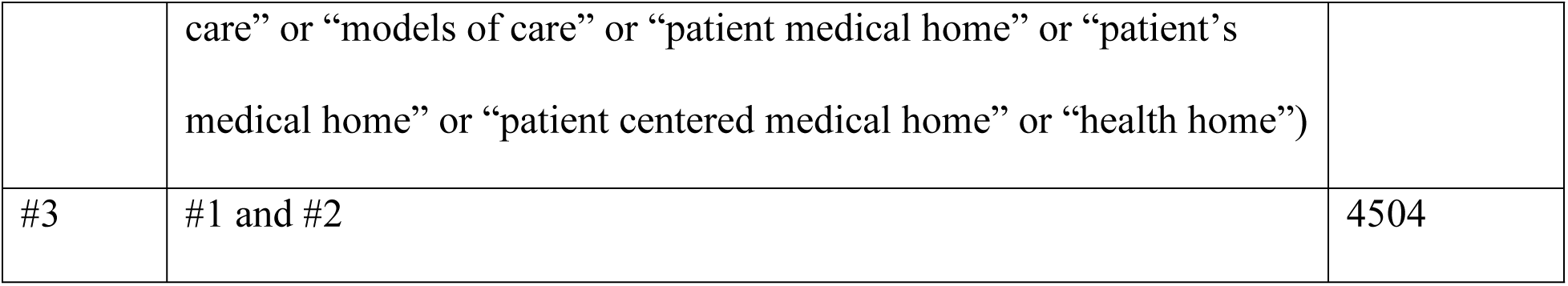
Sample Search Strategy Conducted July 2023 in OVID Medline.

### Study selection

Four members of the research team will take part in the selection process. Each article will have two reviewers independently review the titles and abstracts via COVIDENCE. Screening and eligibility for further review will be determined based on whether the articles presented meet the inclusion criteria. Articles will be included in the full-text review if both reviewers agree that they meet or may meet the eligibility based on the title and abstract. Articles will be excluded if both reviewers agree, based on the title and abstract, that the article does not meet the inclusion criteria. Articles identified as potentially meeting the inclusion criteria will undergo a full-text review, using two independent reviewers to agree on inclusion or exclusion in the scoping review. Any disagreements at either the title and abstract or full-text review phases will be discussed between the two independent reviewers with the goal of reaching a consensus decision. A third reviewer will be consulted to resolve the disagreement if an agreement is not reached. A narrative and flow diagram aligned with the PRISMA-ScR guidance will summarize the study selection process [19].

### Data extraction

Two independent reviewers will extract Data from the articles in the scoping review using a data extraction tool developed and pre-piloted by the review team. Any disagreements arising between the reviewers extracting the data will be resolved through discussion or consultation with an additional reviewer if needed. The data extracted will include details about the study design and methods, characteristics of the primary care model, context, participants, outcomes (if relevant), barriers and facilitators to implementation (if relevant), and other key findings relevant to the review questions.

Table 4 includes a draft extraction form. The draft data extraction tool has been pilot-tested using a preliminary search and revised based on its usability. The data extraction form will be modified as necessary throughout the extraction process from each included article. Any modifications made will be reported in the scoping review. Authors of papers will be contacted to request missing or additional data, where required.

**Table 4.**
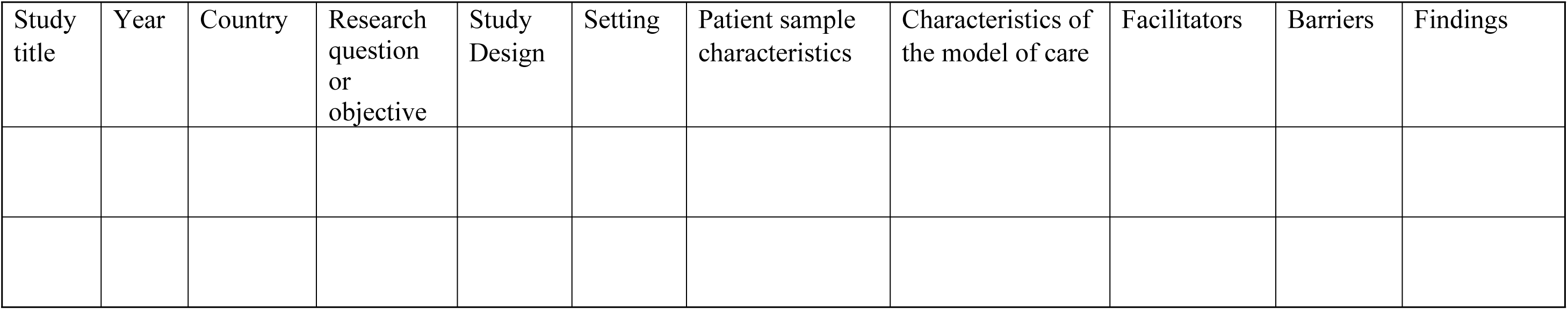
Example Data Extraction Form.

### Data analysis and presentation

For objective 1, the objectives/questions, design, methods, and findings of each included article will be mapped and reported using a table format that includes columns for study design, study objectives/questions, population, outcomes (if relevant), and key findings. A narrative synthesis will accompany the tabulated results. The question/objectives and findings will be critically analyzed to identify the types of evidence available to guide the implementation of models of primary care that include physiotherapists within primary care teams and evidence gaps that need to be addressed through further research. This critical analysis will be reported narratively.

For objective 2, characteristics of the primary care models that integrate physiotherapists within primary care teams will be reported in a table format. The characteristics will be analyzed to identify similarities and differences across the models of care identified and this analysis will be presented using a narrative synthesis to accompany the tabular results.

For objective 3, barriers and facilitators to implementing primary care models that integrate physiotherapists extracted from studies that investigate barriers and facilitators will be analyzed using directed content analysis using the revised Consolidated Framework for Implementation Research (CFIR) [20]. Results will be presented diagrammatically with an accompanying narrative synthesis. This analysis aims to identify factors influencing the implementation of primary care models that integrate physiotherapists within primary care teams [19].

## Discussion

This scoping review will outline the existing literature on primary care models that involve the integration of physiotherapists within primary care teams. The findings will synthesize evidence available to guide the implementation of models of primary care that involve the integration of physiotherapists and highlight gaps in evidence that need to be addressed. The findings will also include a description of the characteristics of the models of care that involve the integration of physiotherapists in primary care, with an analysis of similarities and differences across models of care. Finally, the review will identify barriers and facilitators to implementing models of care that involve the integration of physiotherapists within primary care teams, which is anticipated to provide important evidence to inform the implementation of similar models of care in the future. The findings related to each of the three objectives will provide insight into models of team-based primary care that leverage the expertise of physiotherapists, which is gaining attention worldwide.

## Authors’ contributions

AT and JM developed the project’s concept. All team members contributed equally to developing the protocol, piloting the search strategy, and developing the data extraction sheet. NS coordinated the writing of the first draft of the manuscript, and AT and JM revised the manuscript and provided supervision and support for the project.

## Data Availability

No datasets were generated or analysed during the current study. All relevant data from this study will be made available upon study completion.

## Acknowledgements

We would like to thank Abdul Pullattayil, librarian at Queen’s University, for his advice concerning our search strategy.

## Supporting Information

(Preferred Reporting Items for Systematic Review and Meta-Analysis Protocols)

